# Associations between gas stove usage and childhood asthma symptoms, attacks, and ED visits among children in the 2020 Asthma Call-back Survey

**DOI:** 10.1101/2024.06.04.24308398

**Authors:** Sydney LeSon, Samantha Rosenthal

## Abstract

**Objective:** To examine whether the use of gas stoves in the home is associated with increased asthma severity among children and adolescents ages 0-17 in the US.

**Methods:** Using the 2020 CDC Asthma Call-Back Survey for children, the association between gas stove usage and childhood asthma symptoms, asthma attack or episode, and emergency department visit for asthma was assessed. With a cross-sectional study design, bivariate analyses and multivariable logistic regression were conducted. Survey weights were used in the analyses for US population-based estimates.

**Results:** Children who live in a household that uses gas for cooking or has a gas stove had 1.133 (95% CI: 0.48, 2.68)) times the odds of having an asthma attack or episode within the past 12 months, 9.141 (95% CI: 1.99, 42.06) times the odds of having visited the emergency department or urgent care within the past 12 months, and 1.739 (95% CI: 1.02, 2.95) times the odds of recent symptoms of asthma compared to children who live in a household that does not use gas for cooking or does not have a gas stove, controlling for all confounders. There is an association between the usage of gas stoves and asthma symptoms, asthma attacks/episodes, and ED visits among asthmatic children. Reducing the exposure of gas stove usage should be a consideration in regards to existing and future interventions to prevent childhood asthma and reduce exacerbation of underlying childhood asthma.

## Introduction

Childhood asthma is a chronic lung disease causing a child’s airways to become inflamed when exposed to certain triggers including pollen, catching a cold, or other respiratory infections (1). While childhood asthma is not different from asthma among adults, childhood asthma is the leading cause of emergency department visits, hospitalizations, and missed school days among children in the United States (1). According to the US Environmental Protection Agency, cooking with gas stoves creates nitrogen dioxide, which are irritants to the mucosa of the eyes, nose, throat, and respiratory tract (2). Low level exposure to nitrogen dioxide can increase the risk of respiratory infections in young children, including asthma (2). There is heterogeneous research present regarding the association between indoor gas stove use and childhood asthma. A 2013 meta-analysis of 41 studies on the association between indoor nitrogen oxide and gas cooking on asthma in children concluded that gas cooking increases the risk of asthma (3). This meta-analysis had limitations; notably, all studies included were observational, and there was heterogeneity that existed in the form of participant demographics, study region, proportion of gas cooking, and year of publication between studies (3).

On a global scale, existing literature includes a prospective cohort study regarding asthma and gas stove usage conducted among Puerto Rican youth aged 6-20 years old (4). Questionnaires regarding household characteristics and child’s respiratory health, as well as spirometry were conducted. The study concluded that participants reporting gas stove use had 2.45 times significantly higher odds of asthma compared with those reporting elective stove use (4). Furthermore, a cross-sectional study in 2018 among Australian children under age fourteen associated exposure to gas cooktop stoves with a considerable proportion of childhood asthma burden (5). While these studies show there is research globally on the relation between childhood asthma and gas stove usage, it has become an ongoing discussion in the US that requires additional research to add to the growing, yet conflicting, body of literature.

In early 2023, several news sources began reporting the link between the usage of gas stoves and increased risk of childhood asthma in the United States. A study published in December 2022 calculated that 12.7% of current childhood asthma in the US is attributable to gas stove use (6). Despite this statistic quickly being used by media and political decision makers, the study relied on aggregate data using 27 manuscripts collected through a literature search of peer-reviewed manuscripts published to the most recent meta-analysis (6). Evidently, causal inference cannot be assumed from this data, and a degree of inaccuracy and bias are present - a common setback of a large proportion of studies on asthma and gas stoves revealed by a systematic review through 2022 (7). Inaccuracy stemmed from odds ratios that were not adjusted for or characterized by differences in covariates, which may explain the heterogeneity between studies (8). Furthermore, the key study claimed causation without discussing any bias that stemmed from the lack of external validity (8). With the current discussions regarding gas stoves in homes, it is necessary to establish high quality and generalizable research to support decision-making.

In this study, the association between the use of indoor gas stoves and asthma severity, frequency of asthma attacks, and symptoms among children in the United States will be examined. Using a cross-sectional analysis, indicators of asthma severity, frequency of asthma attacks and symptoms of asthma, are assessed as the outcome. It is hypothesized that gas stove usage currently present in the home is associated with at least one or more of the outcomes of increased symptoms of asthma, higher rate of asthma attack, and increased rates of emergency department or urgent care visits due to asthma.

## Materials and Methods

### Survey Data Description

The Behavioral Risk Factor Surveillance System (BRFSS) is a yearly national, state-based, cross-sectional, random telephone survey developed and monitored by the Center for Disease Control and Prevention (9). The BRFSS is conducted by state health departments using a standardized questionnaire, and is used to collect prevalence data among US residents regarding risk behaviors and preventative health practices (10). To adjust for under-represented groups in the sample to more accurately represent the national population, the BRFSS uses a disproportionate, stratified sampling and includes an iterative proportional fitting weighting method (11). The 2020 BRFSS data had 401,958 individuals who consisted of a random sample of adults aged 18 or older, surveying one per household (11). Some states can elect to include questions regarding children in the survey.

The Asthma Call-back Survey (ACBS) is conducted within 2 weeks after the BRFSS is conducted (12). For states that include children in the BRFSS, if the randomly selected child has ever been diagnosed with asthma, the child is eligible to be surveyed for the child ACBS. The ACBS record included all child variables asked in the BRFSS data in addition to ACBS interview data (12). The ACBS also models the same complex sampling design as the BRFSS, but using child weights. The BRFSS provided demographic data variables for child sex, race/ethnicity, and household income. The ACBS provides information about children with asthma such as demographics, symptoms, healthcare utilization and costs, environmental factors, and asthma management practices. In the 2020 ACBS, 7 states collected child data (CT, GA, MN, NJ, TX, UT, and VT) with a total of 701 participants who completed the child ACBS survey (12). Figure 1 depicts the inclusion and exclusion criteria resulting in the final analytic sample of N = 606 participants. The weighting scheme for both surveys allows for the results to be unbiased and a better representation of the US population. Using the ACBS data, the study used de-identified publicly available data, and therefore did not require the approval of an institutional review board.

**Figure 1.**
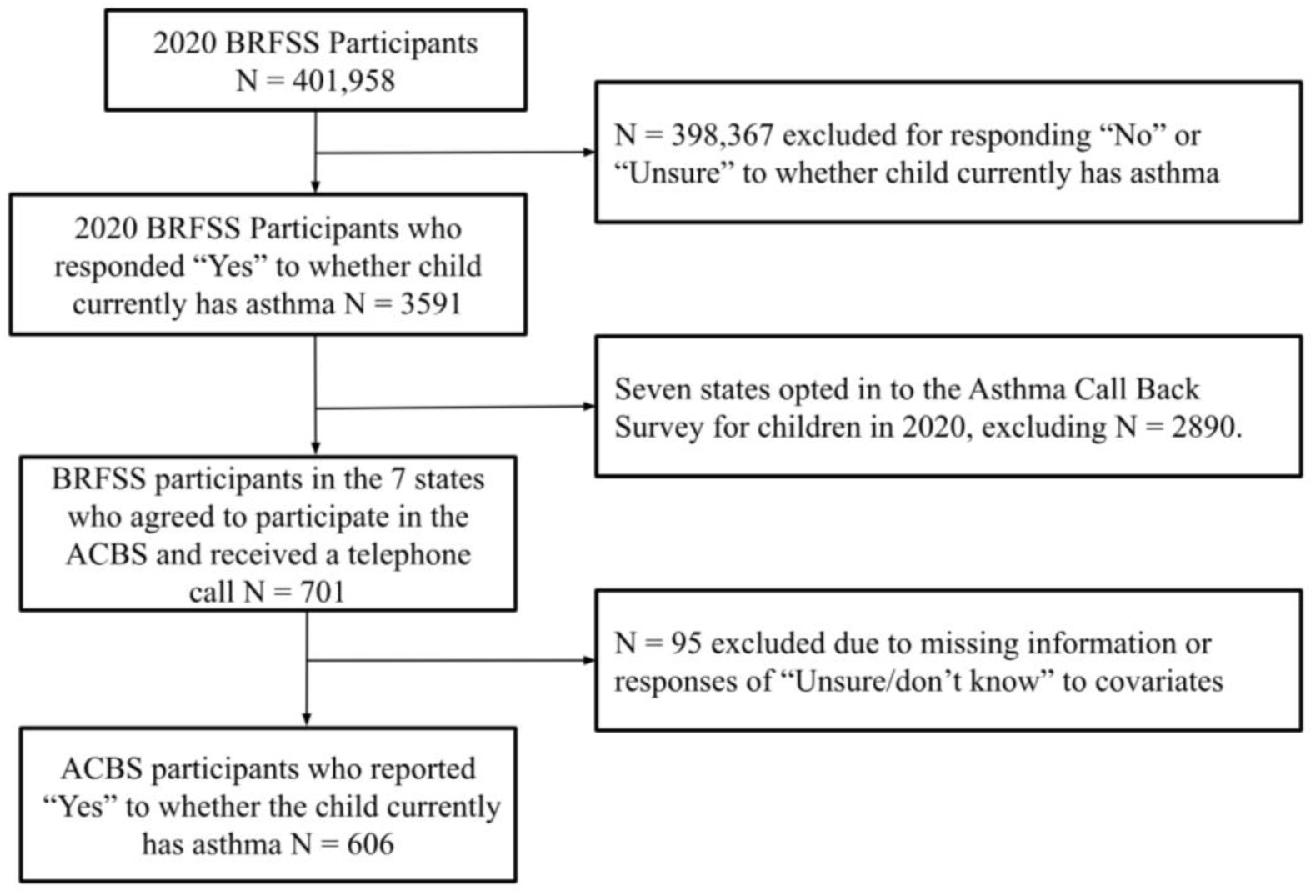
Analytic Sample

### Dependent Variables

The dependent variables (outcomes being assessed) are last occurring symptoms of asthma, asthma attack or asthmatic episode in the last 12 months, and emergency department (ED) or urgent care visit in the last 12 months. Last occurring symptoms of asthma were measured using the survey question “How long has it been since [he/she] last had any symptoms of asthma?”. The symptoms were categorized into a dichotomous variable of less than 1 year ago and 1 year or greater. Asthma attack or asthmatic episode in the last 12 months was measured using the survey question “During the past 12 months, has [he/she] had an episode of asthma or an asthma attack?”. Asthma attack in the last 12 months was categorized into a dichotomous variable of “Yes” and “No”. Emergency department or urgent care visit in the last 12 months was measured using the survey question “During the past 12 months, has [child’s name] had to visit an emergency room or urgent care center because of [ his/her] asthma?”. This variable was categorized into a dichotomous variable of “Yes” and “No”. If any responses were “don’t know/ not sure”, “refused to answer”, or “Question Skipped (symptoms over 12 months ago)”, these observations were excluded in the analysis.

### Independent Variables

The exposure variable of gas stove usage was measured using two questions on the survey. The first question was “Is gas used for cooking in [his/her] home?”. Possible responses were “Yes”, “No”, and “Don’t Know / Not Sure”. The second question was “Are unvented gas logs, unvented gas fireplace, or unvented gas stoves used in [his/her] home?”. Possible responses were “Yes”, “No”, and “Refused”. The exposure variable used these two questions to organize into two categories: “Yes” indicated the respondent answered “Yes” to one or both of the questions, and “No” indicated the respondent answered “No” to both questions. Any responses of “don’t know/not sure” and “refused” were excluded from the analysis.

### Confounders

Potential confounders in the association between gas stoves and indicators of asthma included demographic, socioeconomic, and environmental factors. Individual characteristics of age, sex, race/ethnicity, and household income were included based on prior literature indicating race, sex, and socioeconomic status disparities in the prevalence and burden of asthma (13). The American Lung association states that pet allergens are a known risk factor for asthma flare-ups and more severe asthma in children if pets are present in the home (14). In a cross-sectional study among US children, children living in homes that used ventilation when using gas stoves had lower odds of asthma compared to homes that never used ventilation (15). As such, related environmental variables such as pets in the household and use of kitchen exhaust fans were included in this study.

Age was categorized into five discrete intervals (< 1, 1-2, 3-5, 6-10, and 11-17 years old). Sex was categorized as male and female. Household income was determined by respondent’s self-reported annual income and categorized into 3 discrete variables (< $25000, $25000 – $49999, and ≥$50000). Race was categorized into four groups: White, Black, Hispanic, and Other. The category of Other encompasses responses of American Indian or Alaskan Native, Asian, Multiracial, and Other. Pets in the home were categorized into a dichotomous variable “Yes” and “No” based on whether the child’s home had any pets that spend time indoors. Kitchen fan usage was categorized into a dichotomous variable “Yes” and “No” based on whether an exhaust fan that vents to the outside was used regularly when cooking. All responses that were missing, or responded “Don’t know/unsure”, or “refused” were excluded from the analysis.

### Analysis

The following statistical analyses were performed using SAS statistical software version 9.4 (16). Table 1 and Table 2 have unweighted frequency (N), weighted frequency (n), and weighted percentages (%) reported. Bivariate analyses were conducted to examine relationships between the selected characteristics, gas stove usage, and the asthma severity indicator variables. Specifically, Rao-Scott chi-squared tests were calculated, a design-adjusted version of the Pearson chi-square test (17). Two-sided *p-*values were collected to measure the statistical significance of the association between gas stoves and each asthma outcome. A *p-*value < 0.05 was considered statistically significant. Separate models were run for each asthma outcome variable (asthma symptoms, recent asthma attack/episode, and asthma-related ED or urgent care visit).

**Table 1.** Characteristics of asthmatic children in the 2020 Asthma Call-Back Survey by gas stove usage.

| **Selected Characteristic** | **Gas stoves are not in home or gas is not used for cooking**<br>**(N = 314 (50.28%))**<br>**[n (Weighted %)]** | **Gas stoves are in home or gas is used for cooking**<br>**(N = 292 (49.72%))**<br>**[n (Weighted %)]** | **P-value<sup>a</sup>** |
| --- | --- | --- | --- |
| Age |  |  | 0.7940 |
| <1 | 44 (44.31) | 54 (55.68) |  |
| 1-2 | 78 (51.30) | 83 (48.70) |  |
| 3-5 | 106 (51.65) | 76 (48.34) |  |
| 6-10 | 62 (50.99) | 65 (49.01) |  |
| 11-17 | 24 (72.20) | 14 (27.80) |  |
| Gender |  |  | 0.9410 |
| Male | 178 (49.99) | 172 (50.01) |  |
| Female | 136 (50.75) | 120 (49.25) |  |
| Household Income |  |  | <.0001 |
| < \$25,000 | 83 (65.64) | 50 (35.36) | |
| \$25,000 – \$49,999 | 62 (68.19) | 32 (31.81) | |
| ≥ \$50,000 | 169 (35.64) | 210 (64.36) | |
| Race and Ethnicity |  |  | 0.2990 |
| White | 233 (44.58) | 216 (55.41) |  |
| Black | 41 (59.98) | 33 (40.02) |  |
| Hispanic | 16 (70.55) | 15 (29.45) |  |
| Other | 24 (42.52) | 28 (57.48) |  |
| Pets |  |  | 0.8523 |
| Yes | 205 (51.16) | 200 (48.84) |  |
| No | 109 (49.40) | 92 (50.60) |  |
| Kitchen Fan Usage |  |  | 0.2227 |
| Yes | 218 (52.02) | 210 (47.98 ) |  |
| No | 96 (42.85) | 82 (57.15) |  |
| Last occurring Symptoms |  |  | 0.3685 |
| < 1 year | 170 (46.84) | 164 (53.16) |  |
| ≥ 1yr | 135 (55.61) | 122 (44.39) |  |
| Asthma Attack or Episode in past 12 months |  |  | 0.8083 |
| Yes | 106 (45.63) | 97 (54.37) |  |
| No | 70 (47.32) | 69 (52.68) |  |
| ED Visit in last 12 months |  |  | 0.0071 |
| Yes | 20 (22.20) | 34 (77.80) |  |
| No | 294 (54.5) | 257 (45.47) |  |

<sup>a</sup>p-values were calculated using the Rao-Scott Chi-Square test

**Table 2.** Characteristics of asthmatic children in the 2020 Asthma Call-Back Survey by asthma attack and emergency department visit in the past 12 months.

|  | **Last occurring Symptoms in 12 months (N = 334 (57.0%))**<br>**[n (Weighted %)]** | **P-value<sup>a</sup>** | **Asthma attack in past 12 months (N = 203 (55.93%))**<br>**[n (Weighted %)]** | **P-value<sup>a</sup>** | **ED or Urgent Care visit in past 12 months (N = 54 (13.00%))**<br>**[n (Weighted %)]** | **P-value<sup>a</sup>** |
| --- | --- | --- | --- | --- | --- | --- |
| Gas Stoves |  | 0.3685 |  | 0.8083 |  | 0.0071 |
| Yes | 164 (30.30) |  | 97 (30.41) |  | 34 (10.11) |  |
| No | 170 (26.70) |  | 106 (25.52) |  | 20 (2.89) |  |
| Age |  | 0.0171 |  | 0.0696 |  | 0.3594 |
| >1 | 45 (10.12) |  | 29 (6.05) |  | 13 (4.2) |  |
| 1-2 | 81 (16.77) |  | 46 (17.62) |  | 18 (4.23) |  |
| 3-5 | 101 (21.32) |  | 69 (23.02) |  | 17 (4.03) |  |
| 6-10 | 82 (7.03) |  | 47 (7.84) |  | 5 (0.51) |  |
| 11-17 | 25 (1.75) |  | 12 (1.39) |  | 1 (0.021) |  |
| Gender |  | 0.2165 |  | 0.0341 |  | 0.0008 |
| Male | 185 (37.40) |  | 111 (33.68) |  | 36 (10.68) |  |
| Female | 149 (19.60) |  | 92 (22.25) |  | 18 (2.32) |  |
| Household Income |  | 0.2990 |  | 0.3911 |  | 0.1827 |
| < \$25,000 | 80 (21.66) | | 43 (19.57) | | 14 (5.69) | |
| \$25,000 – \$50,000 | 52 (9.07) | | 34 (8.43) | | 13 (0.94) | |
| ≥ \$50,000 | 202 (26.27) | | 126 (27.93) | | 27 (6.37) | |
| Race and Ethnicity |  | 0.9173 |  | <.0001 |  | 0.1420 |
| White | 246 (31.72) |  | 159 (34.60) |  | 32 (5.86) |  |
| Black | 48 (16.80) |  | 23 (9.21) |  | 11 (5.84) |  |
| Hispanic | 16 (2.19) |  | 8 (2.31) |  | 4 (0.33) |  |
| Other | 24 (6.29) |  | 13 (9.81) |  | 7 (0.97) |  |

<sup>a</sup>p-values were calculated using the Rao-Scott Chi-Square test

Both the BRFSS and child ACBS weighting included design weighting and iterative proportional fitting (12). Data was analyzed using appropriate variables for stratification, clustering, and sample weight to account for the complex sampling design of the survey data. Weighted data was used to estimate the population parameters and account for any non-response and non-coverage that was present in the BRFSS and child ACBS interview (12).

Table 3 reports all crude and adjusted odds ratios for each asthma outcome. Simple logistic regression was conducted to determine crude odds ratios. Adjusted odds ratios were calculated from multivariable logistic regression models that adjusted for all potential confounders.

**Table 3.** Unadjusted and adjusted odds of asthma symptoms, asthma attacks, and emergency department visits among children in the 2020 ACBS.

|  | **Last occurring Asthma Symptoms<sup>a</sup>**<br>**(N = 591)** |  | **Asthma Attack or Episode within in past 12 months<sup>a</sup>**<br>**(N= 203)** |  | **Emergency Department Visit within the last 12 months<sup>a</sup>**<br>**(N= 54)** |  |
| --- | --- | --- | --- | --- | --- | --- |
|  | Crude OR<br>(95% CI) | Adjusted OR<sup>b</sup><br>(95% CI) | Crude OR<br>(95% CI) | Adjusted OR<sup>b</sup><br>(95% CI) | Crude OR<br>(95% CI) | Adjusted OR<sup>b</sup><br>(95% CI) |
| Gas Stove Usage |  |  |  |  |  |  |
| Yes (1 vs 2) | 1.422<br>(0.649, 3.116) | 2.03<br>(1.04, 3.946) | 1.07<br>(0.61, 1.88) | 1.037<br>(0.498, 2.16) | 4.201<br>(1.38, 12.78)) | 11.04<br>(2.349, 51.88) |

Crude and adjusted odds ratios were weighted using survey logistic and ACBS weighting variables (\_psu, \_ststr, cllcpwt\_f).
<sup>a</sup>Confidence intervals calculated using Wald test.
<sup>b</sup>Adjusted odds ratios included all confounding variables (age, sex, race, annual household income, household pets, and usage of kitchen fan)

## Results

The total number of participants who responded to the 2020 Child ACBS, thus eligible for inclusion, was 701. After removing observations of “Don’t Know” or “Refused” from the variables of gas stove usage, and asthma indicators of symptoms, frequency of attack, and ED visit for asthma, 700 observations remained. Removing observations of “Don’t Know” or “Refused” from the remaining covariates resulted in 672 observations. Finally, removing any observations with missing values due to nonresponse resulted in a total of 606 survey responses that comprised the analytical sample for the final analysis.

Of the 606 participants, 292 participants (49.72%) reported using gas for cooking or having an unvented gas stove in their home. As indicated in Table 1, children of the participants surveyed were predominantly white (74.1%), male (57.8%), and came from households with annual income greater than $50,000 (62.5%). Of the environmental factors, the households with pets and who used kitchen fans were fairly equal between homes with gas stove usage compared to no gas stove usage (48.8% with pets and 47.9% using kitchen fans in homes with gas stove usage). Additionally, there was a higher proportion of children who experienced asthma symptoms within the last year (53.5%) compared to children who experienced asthma symptoms over 1 year ago (46.5%) in households both with and without gas stove usage. Among households with gas stove usage, children had a higher percentage of an ED visit in the last 12 months (77.8%) compared to children in households without gas stove usage.

From Table 2, children who were aged 5 or under (88.9%) were more likely than children aged 6 and older to have any emergency department or urgent care visit in the past 12 months. Additionally, children aged 5 or under were more likely to have an asthma attack or episode in the past 12 months (70.9%) compared to children aged 6 and older. For children with an asthma attack and ED visit in the last 12 months, there was a significant difference between males and females (asthma attack *p*-value = 0.03, ED visit *p*-value = 0.0008). There was also a significant difference in ED visits for asthma between homes with gas stove usage and homes without gas stove usage (*p*-value = 0.0071).

In the models shown in Table 3, children who live in a household that uses gas for cooking or has a gas stove had 1.07 (95% CI: 0.61, 1.88) times the odds of having an asthma attack or episode within the past 12 months compared to children who live in a household that does not use gas for cooking or does not have a gas stove; yet, when controlling for all other variables, the effect remained significant (adjusted OR 1.133 (95% CI: 0.48, 2.68)). Children who live in a household that uses gas for cooking or has a gas stove had 4.201 (95% CI: 1.38, 12.78) times the odds of having visited the emergency department or urgent care within the past 12 months compared to children who live in a household that does not use gas for cooking or does not have a gas stove; when controlling for all other variables, the effect remained significant (adjusted OR 9.141 (95% CI: 1.99, 42.06)). Children who live in a household that uses gas for cooking or has a gas stove had 1.37 (95% CI: 0.70, 2.70) times the odds of recent symptoms of asthma compared to children who live in a household that does not use gas for cooking or does not have a gas stove; controlling for all other variables, the effect remained significant (adjusted OR 1.739 (95% CI: 1.02, 2.95)).

## Discussion

This study expands upon existing literature to observe asthma outcomes currently bereft in the literature through observing asthma symptoms, asthma episodes and attacks, as well as ED visits among children. Among children with asthma, children who live in homes with gas stove usage were significantly more likely to have an ED visit in the past 12 months compared to children living in homes without gas stove usage. Children with asthma aged 0-10 had an increased likelihood to have symptoms occurring within the last 12 months compared to children aged 11-17, showing higher prevalence of asthma symptoms among younger children. Children living in homes that used gas stoves had a higher odds of having asthma symptoms and an asthma attack occur in the last year compared to children living in homes that do not use gas stoves, adjusting for all confounders. Our findings suggest that children with asthma who live in homes with a gas stove were more likely to have asthma symptoms, asthma attack or episode, and an ED visit in the past year.

A review of existing literature has highlighted epidemiological flaws within the design and analysis of the studies. The highly cited statistic by Gruenwald et al. states that gas stove emissions cause 13% of childhood asthma in the United States (6). Within the study, Gruenwald et al. calculates population attributable fractions to prevalence estimates and gas stove data from an existing 2013 meta-analysis by Lin et al (6). Within this meta-analysis, Lin et al. concludes from calculated odds ratios that children living in a home with gas cooking have a 42% increased risk of having current asthma (3). Limitations within this meta-analysis include unadjusted confounders between associations, as well as differences in timing, populations, settings, and location of data collected, that question the generalizability, reliability, and validity of recent literature (8). Both papers essentially conduct statistical associations. Thus, the causal conclusions that these papers generate were inappropriate due to the lack of data supporting valid causal inferences and any true causal analysis methods (8).

Following this meta-analysis, Li et al. conducted the first systematic review on gas cooking and asthma or wheeze in children to evaluate study heterogeneity and study quality (7). The systematic review identified sources of biases and inaccuracy due to self-reporting measures, lack of adjustment of confounders, and unestablished temporality (7). Three studies in the review compared the prevalence of gas cooking exposure between children with versus without asthma, and three studies compared the prevalence of asthma between children in homes using gas vs electricity for cooking; all of these studies reported statistical insignificance but did not adjust for any potential confounders (7). In the previous studies within Li et al.’s systematic review that did account for the adjustment of confounding in their analysis, twenty studies similarly relied on self-reported information. The prevalence of gas cooking exposure varied between studies; our study reported a gas stove usage prevalence of 49.72% while other studies ranged from 1.3% to 94.09% globally (7). With respect to the outcome of these studies, newly diagnosed asthma, ever diagnosed asthma, and asthma exacerbation were included (7).

As previous studies may have overstated their results, our study expands upon the current body of literature regarding gas stoves and asthma outcomes through providing transparent and appropriate conclusions. The relative measures of association calculated, both crude and adjusted measures, provide clear directions of association regarding gas stoves and each asthma outcome. This study assists in addressing current conflicting bodies of evidence with appropriate epidemiological inferences.

There are a few limitations to this study. One limitation arises from the cross-sectional study design; temporality cannot be established, and thus causality cannot be confirmed with these associations. Moreover, gas stove usage information was determined by a “yes” or “no” response to whether gas was used or present while symptoms, asthma attack or episode, and emergency department or urgent care visit for asthma were limited to the past 12 months. Due to the cross-sectional nature and differences in timing of the question asked, it is not possible to determine whether these measures of current asthma occurred exactly before, after, or concurrently with gas stove usage in the family home.

In regards to the data collection, the median response rate for the 2020 BRFSS was 47.9%, and the response rate of the 2020 Child ACBS was 84.34% (18, 19). Although the BRFSS has limitations due to the lower response rate, the survey is considered to be one of the few available nationally representative health surveys with robust weighting to account for coverage and non-response, allowing for population-based estimates (18). The child ACBS had a higher response rate and is essential in addressing childhood asthma. The sample size is limited by which states decide to opt in to conducting the ACBS for children. Furthermore, a significant limitation of the study is that the data collected from the ACBS was based on self-reported responses over the phone. Therefore, participant responses in the survey are subject to social desirability bias as well as recall bias. Potential residual confounding may exist from other unmeasured comorbidities.

More specifically, there is uncertain validity of self-reported asthma status for children in the BRFSS. The BRFSS asks “Has a doctor, nurse, or other health professional ever said that the child has asthma?” (20). The follow up question is “Does the child still have asthma?” (20). Despite these questions, there is potential that the clinical diagnosis or respondent’s recall of the diagnosis is inaccurate, excluding any true asthmatic children from the sample. A 1998 review of asthma questionnaires reported a mean sensitivity of 68% and mean specificity of 94% when self-reported asthma was compared with clinical diagnosis of asthma (21). Based on this review, the higher specificity indicates that individuals who do not have a clinical diagnosis of asthma have higher rates of self-reporting no asthma.

Despite these limitations, this study included a large sample of asthmatic children using a US nationally representative sample. The BRFSS remains the world’s largest telephone survey which includes the most comprehensive source of asthma surveillance for children (18). Furthermore, it provided new empirical evidence regarding the relationship between gas stove usage and asthma symptoms, episodes, and related ED visits generalizable to children with asthma in the US.

The results suggest that gas stove usage should be considered in asthma prevention interventions among children. Possible intervention includes replacing stoves using gas with electric stoves or any cleaner alternative. If a child currently has asthma, the gas stove should be removed from the home. In addition, strategies for reducing exposure to gas stoves and appliances should be created and communicated to parents of children who currently have asthma or may be at risk (5). Health policy should be considered to eliminate gas stove usage in homes to reduce increased risk of current childhood asthma. In May 2023, New York was the first state to pass legislative action to require new buildings to ban gas stoves and furnaces starting in 2026 (22). Current policy debate exists on the federal level regarding the ban of gas stove usage for energy saving and curbing climate change (23). Legislation to ban gas stove usage may be beneficial to alleviate existing asthma symptoms, attacks, and ED visits in children.

This study used relative measures of association as a method to estimate potential health impact if gas stove usage was eliminated in homes with children with current asthma. Moreover, the relative measurements calculated will only hold if gas stove usage is causally related to these indicators of current asthma (symptoms, attack or episode, and/or ED visit in the past 12 months) among children. Thus, additional research should continue to establish temporality and causality such as through longitudinal study designs to better understand effects of gas stove usage on current childhood asthma.

## Conclusion

There is an association between the usage of gas stoves and asthma symptoms, asthma attacks/episodes, and ED visits among asthmatic children, and as such increased attention should be focused on this relationship specifically in regards to reducing these outcomes. Although current debate exists in regards to gas stove policies at the state and federal level, policy should consider providing alternatives to gas stove usage in homes as an intervention to alleviate exacerbation of childhood asthma. This study provides novel insight in regards to asthma symptoms, asthma attacks, and ED visits for asthma in the last year which are specific outcomes absent in the literature. Additional longitudinal studies should be conducted to confirm external validity and causality. Reducing the exposure of gas stove usage should be a consideration in regards to existing and future interventions to prevent childhood asthma and reduce exacerbation of underlying childhood asthma.

## Data Availability

All data utilized in the present study are available online at https://www.cdc.gov/brfss/acbs/2020_documentation.html

